# Neutralizing Antibody Responses in COVID-19 Convalescent Sera

**DOI:** 10.1101/2020.07.10.20150557

**Authors:** William T. Lee, Roxanne C. Girardin, Alan P. Dupuis, Karen E. Kulas, Anne F. Payne, Susan J. Wong, Suzanne Arinsburg, Freddy T. Nguyen, Damodara Rao Mendu, Adolfo Firpo-Betancourt, Jeffrey Jhang, Ania Wajnberg, Florian Krammer, Carlos Cordon-Cardo, Sherlita Amler, Marisa Montecalvo, Brad Hutton, Jill Taylor, Kathleen A. McDonough

**Author notes:** Corresponding Author: William T. Lee. Authors, other than those affiliated with Mount Sinai Hospital, as above, do not have a commercial or other association that might pose a conflict of interest. This work has not been presented elsewhere. Address correspondence and reprint requests to: Dr. William Lee, The Wadsworth Center/NYSDOH, David Axelrod Institute, 120 New Scotland Avenue, Albany, NY 12208, (email).

## Abstract

Passive transfer of antibodies from COVID-19 convalescent patients is being used as an experimental treatment for eligible patients with SARS-CoV-2 infections. The United States Food and Drug Administration’s (FDA) guidelines for convalescent plasma recommends target antibody titers of 160. We evaluated SARS-CoV-2 neutralizing antibodies in sera from recovered COVID-19 patients using plaque reduction neutralization tests (PRNT) at low (PRNT50) and high (PRNT90) stringency thresholds. We found that neutralizing activity increased with time post symptom onset (PSO), reaching a peak at 31-35 days PSO. At this point, the number of sera having neutralizing titers of at least 160 was ∼93% (PRNT50) and ∼54% (PRNT90). Sera with high SARS-CoV-2 antibody levels (>960 ELISA titers) showed maximal activity, but not all high titer sera contained neutralizing antibody at FDA recommended levels, particularly at high stringency. These results underscore the value of serum characterization for neutralization activity.

## Introduction

The United States has been profoundly impacted by Coronavirus Disease 2019 (COVID-19) since the movement of Severe Acute Respiratory Syndrome coronavirus 2 (SARS-CoV-2) viral infections out of China and across the globe in early 2020. As of May 14, 2020, the United States had 1,390,764 confirmed COVID-19 cases and more than 84,000 COVID-19-related deaths [1] [2]. The first documented COVID-19 case in New York was reported on March 1, 2020. New York State currently accounts for the largest portion of cases (340,661 with 27,477 deaths) in the United States [1] [3].

Infection with SARS-CoV-2 can be severe, especially among at risk populations (e.g., the elderly, especially with pre-existing co-morbidities) [4]. Nonetheless, up to 80% of infections are thought to be mild or asymptomatic [5]. Humoral, or antibody (Ab)- mediated, immunity is thought to protect an individual from viral infection by interfering with virus-host cell interactions required for viral entry and replication. Vaccinated or previously infected individuals with virus-specific Abs can also help prevent new infections through herd immunity [6]. However, the duration and degree to which asymptomatic infection or recovery from COVID-19 disease confers prolonged immunity from reinfection with SARS-CoV-2 is unclear, even among individuals who produce virus-specific antibodies [7] [8]. Consequently, there a great interest in gaining a better understanding of Ab responses to SARS-CoV-2 and the serological tests that measure them [9] [10].

Due to the lack of effective treatments, current COVID-19 outbreak management emphasizes social distancing, expanded diagnostic testing and isolation of known cases and quarantine of close contacts. The resulting economic and societal disruption exacerbates the public health impact of COVID-19, and there is an urgent need to define correlates of immunological status to identify individuals who may have protective immunity. Passive transfer of Ab from convalescent COVID-19 patients is being used as treatment of seriously ill COVID-19 patients, although its efficacy has not yet been rigorously evaluated. Recovered COVID-19 patients who have generated robust Ab responses to SARS-CoV-2 are being sought to fill a growing need for plasma donors to support this therapy. A lack of standardization or information about the relative neutralizing capacity of Abs from convalescent donors complicates implementation and evaluation of plasma therapy protocols for COVID-19. The U.S. Food and Drug Administration (FDA) has issued a recommendation that the titer of neutralizing Abs in convalescent plasma should be at least 160, but allows that an 80 titer is acceptable in the absence of a better match [11]. However, this guidance does not specify the level of virus neutralization that should be achieved at these titers or how to measure it.

Several approaches to measuring relative levels of neutralizing antibody exist, including plaque reduction neutralization tests (PRNT) and microneutralization (MN) [12] [13]. Here we chose the PRNT, which measures the ability of Ab in serially diluted sera or plasma to decrease virus infection of cultured cells at a minimum threshold (e.g., 50% or 90%). PRNT is a relatively low throughput assay because it takes several days for development of virus plaques, which are the units of measurement. MN assays can provide a higher throughput alternative, but the need for BSL3 containment during SARS-CoV-2 culture remains a barrier. As a result of these limitations, determination of Ab neutralization activity has not been routinely adopted by many COVID-19 plasma donor screening programs; instead donors are chosen based on Ab levels or time since clinical recovery from PCR confirmed disease.

The purpose of this study was to measure SARS-CoV-2 specific Abs in the sera of COVID-19 convalescent individuals to characterize the humoral immune response in these individuals, and correlate this response to Ab neutralizing activity, and evaluate neutralizing activity in the context of FDA recommended guidelines for selection of convalescent plasma donors

## Methods

### Study Participants

Specimens were collected with informed consent obtained from individuals in accordance with guidelines of Mount Sinai Hospital (MSH) and the Westchester County Department of Health (WCDH). Testing at the Wadsworth Center was done under a declared Public Health Emergency with a waiver from the NYSDOH Institutional Review Board.

#### Study Set 1

Sera were obtained from individuals who had recovered from COVID-19 during a recruitment effort to identify plasma donors for passive Ab transfer therapy. Seropositivity assessment on these specimens was done by enzyme-linked immunosorbent assay (ELISA) at MSH Laboratory [14, 15]. A sliding recruitment was done to increase the selection of recovered COVID-19 patients at later stages of convalescence. Of the 3277 people who entered the study by April 12, 2020, 227 had a positive SARS-CoV-2 PCR test at MSH, Labcorp, or the New York City Department of Health and Mental Hygiene, and 621 had a self-reported positive PCR result. An additional 1423 had antibodies reactive in the ELISA at 1:50 dilution and were considered “true” COVID-19 recovered individuals.

A progressive approach was used to define eligibility for the donor candidate screening in this cohort. For the first 3 days of the study, the Ab screening criteria included patients who were at least 10 days beyond the onset of symptoms or diagnostic PCR test and asymptomatic for 3 days. With each subsequent week of the study, selected patients were deeper into the convalescent phase, where increased Ab responses would be expected based on other viral infections [16] [17]. By the third week, all patients had to be 21 or more days from the start of infection, and 14 or more days from full recovery.

#### Study Set 2

To specifically determine antibody levels and neutralizing antibodies in individuals well-removed from the onset of symptoms, sera from 149 healthy, recovered COVID-19 individuals from Westchester County were screened as potential donors of passive Ab therapy. The WCDH recruited individuals as potential plasma donors if they had confirmed positive SARS-CoV-2 PCR results at least 21 days before the date of serum collection and were symptom free for at least 14 days. Ascertainment of symptom free status was done by interviewing the individual at the time the arrangements were made for phlebotomy. Antibody testing for the Westchester cohort was done at the Wadsworth Center using a microsphere immunoassay (MIA).

### Immunoassays

#### ELISA

The initial description of this FDA EUA test and methodological details are described elsewhere [14] [15]. This ELISA detects Abs that are reactive with the SARS-CoV-2 receptor binding domain (RBD) and the entire spike protein ectodomain. For most of the sera described in this report, specimens were screened initially at a 1:50 dilution using the SARS-CoV-2 RBD as the target antigen and specific IgG was detected. The endpoint titers of the “presumptive screen positive” sera were then determined by ELISA using the whole recombinant spike ectodomain as the target antigen.

#### MIA

The NY SARS-CoV-MIA is a suspension phase assay using the Luminex platform. This is an FDA EUA test and the details and performance characteristics are described elsewhere [18]. For all specimens included in this study, the MIA measured reactivity to the SARS-CoV-1 nucleoprotein (N) antigen that had been coupled to polystyrene microspheres. Total antigen-specific immunoglobulin was measured using a biotinylated goat anti-human Ig (reactive with IgM, IgA, and IgG), followed by detection with phycoerythrin-conjugated streptavidin. Anti-SARS-CoV-2 N Ab are strongly identified due to extensive amino acid identity between SARS-CoV-1 and SARS-CoV-2 (∼90%), and ongoing studies show that the substitution of the SARS-CoV-2 N protein provides essentially identical results (WTL, unpublished observations). Many of the sera assessed in this study were also evaluated using the SARS-CoV-2 RBD antigen in the MIA, multiplexed in conjunction with the N protein. With some exceptions, both antigens were reactive with the same sera. For the MIA assay positivity is defined as 6 standard deviations (SD) above the mean Median Fluorescence Intensity (MFI) of the signal from 90 pre-2019 blood donor sera from presumed healthy individuals. The high cutoff maximizes specificity (99%) to identify sera with moderate to high amounts of SARS-CoV-2-reactive antibodies and excludes potential cross-reactivity with other respiratory viruses. Results for which the MFI signal falls between 3 SD and 6 SD above the mean MFI of normal sera are considered “Indeterminate”.

#### PRNT

This assay has been previously described and is considered the classical standard for detection of virus-specific Abs based on their ability to neutralize their cognate viral infections [13] [19] [20]. For the detection of SARS-CoV-2 neutralizing Abs, 100 ul of 2-fold serially diluted test sera were mixed with 100 ul of 200 plaque forming units (PFUs) of SARS-CoV-2, isolate USA-WA1/2020 (BEI Resources, NR-52281) and incubated at 37°C in an incubator with 5% CO2 for one hour. 100 ul of virus:serum mixture was added to Vero E6 cells (C1008, ATCC CRL-1586) and adsorption proceeded at 37°C in an incubator with 5% CO2 for one hour, after which a 0.6% agar overlay prepared in cell culture maintenance medium (Eagle’s Minimal Essential Medium, 2% heat-inactivated fetal bovine serum, 100 µg/ml Penicillin G, 100 U/ml Streptomycin) was applied. At two days post-infection, a second agar overlay containing 0.2% Neutral red was applied, and the number of plaques in each sample were recorded after an additional 2 days of incubation. The inverse of the highest dilutions of sera providing 50% (PRNT50) or 90% (PRNT90) viral plaque reduction relative to virus-only infection was reported as the titer.

## Results

### Characterization of tested specimens

For this study period, 3277 specimens from convalescent COVID-19 individuals were screened for SARS-CoV-2 RBD Ab reactivity using the ELISA assay at MSH. Of these specimens, 2398 were also tested for Abs at the Wadsworth Center using the MIA. An additional 171 serum specimens, submitted directly to the Wadsworth Center from the WCDH, were tested for the presence of Abs to the SARS CoV N protein. For the specimens tested at both MSH and the Wadsworth Center, the two assays (ELISA and MIA, respectively) showed 79.3% agreement (Supplementary Table 1). If the MIA indeterminates (3 SD) were also counted as reactive, the agreement increased to 87.6%. Inclusion of RBD with the N protein in the MIA resulted in agreement levels of 89.7.2% (6 SD) and 90.6% (3 SD). A direct comparison of the values provided by the two tests showed the strongest correlation between the MIA RBD protein and the ELISA spike protein (Supplementary Figure 1).

**Table 1.**
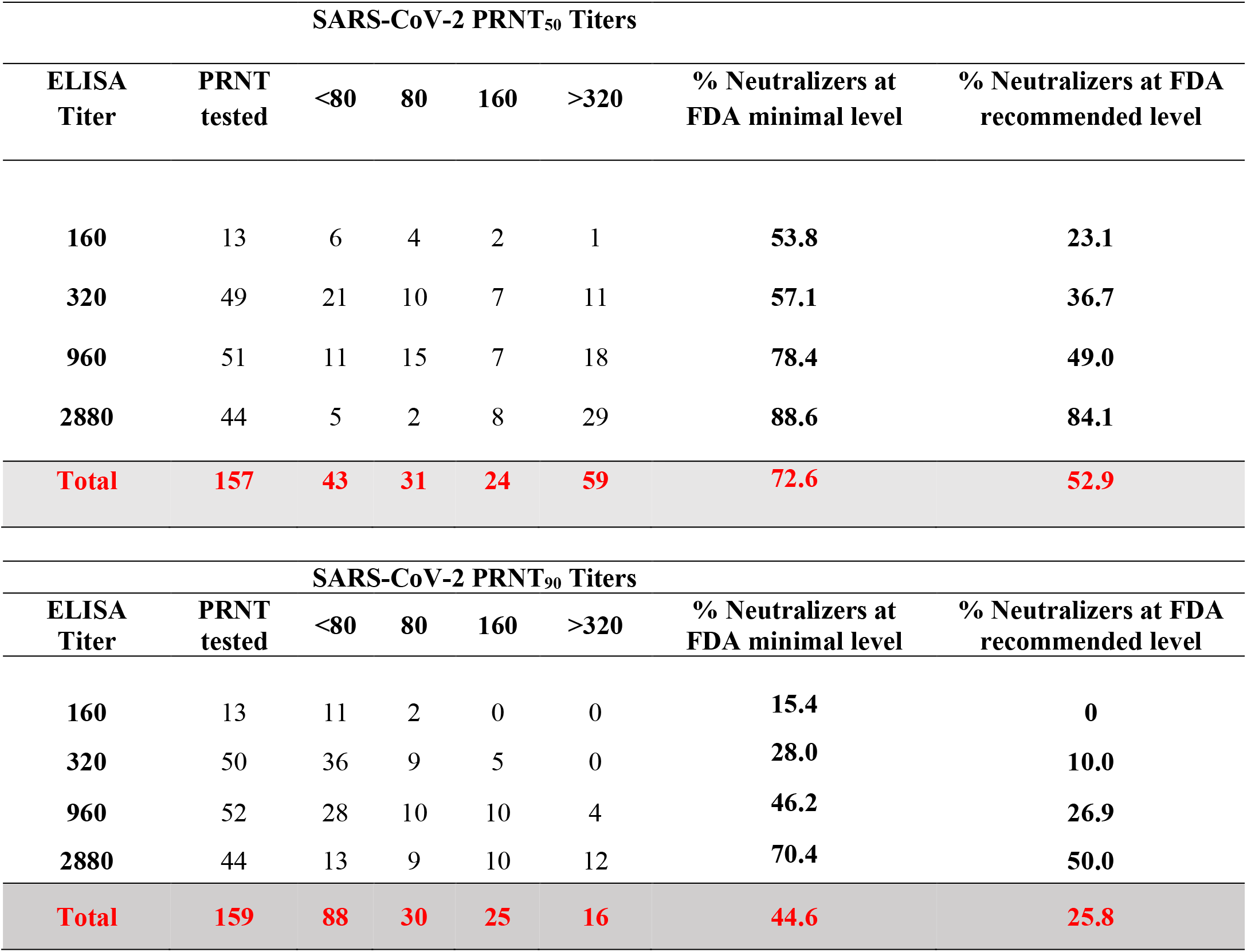
Relationship between SARS-CoV Neutralizing Abs and ELISA Titer.

| SARS-CoV-2 PRNT<sub>50</sub> Titers |  |  |  |  |  |  |  |
| --- | --- | --- | --- | --- | --- | --- | --- |
| ELISA Titer | PRNT tested | <80 | 80 | 160 | >320 | % Neutralizers at FDA minimal level | % Neutralizers at FDA recommended level |
| 160 | 13 | 6 | 4 | 2 | 1 | 53.8 | 23.1 |
| 320 | 49 | 21 | 10 | 7 | 11 | 57.1 | 36.7 |
| 960 | 51 | 11 | 15 | 7 | 18 | 78.4 | 49.0 |
| 2880 | 44 | 5 | 2 | 8 | 29 | 88.6 | 84.1 |
| **Total** | **157** | **43** | **31** | **24** | **59** | **72.6** | **52.9** |

| SARS-CoV-2 PRNT<sub>90</sub> Titers |  |  |  |  |  |  |  |
| --- | --- | --- | --- | --- | --- | --- | --- |
| ELISA Titer | PRNT tested | <80 | 80 | 160 | >320 | % Neutralizers at FDA minimal level | % Neutralizers at FDA recommended level |
| 160 | 13 | 11 | 2 | 0 | 0 | 15.4 | 0 |
| 320 | 50 | 36 | 9 | 5 | 0 | 28.0 | 10.0 |
| 960 | 52 | 28 | 10 | 10 | 4 | 46.2 | 26.9 |
| 2880 | 44 | 13 | 9 | 10 | 12 | 70.4 | 50.0 |
| **Total** | **159** | **88** | **30** | **25** | **16** | **44.6** | **25.8** |

**Figure 1.**
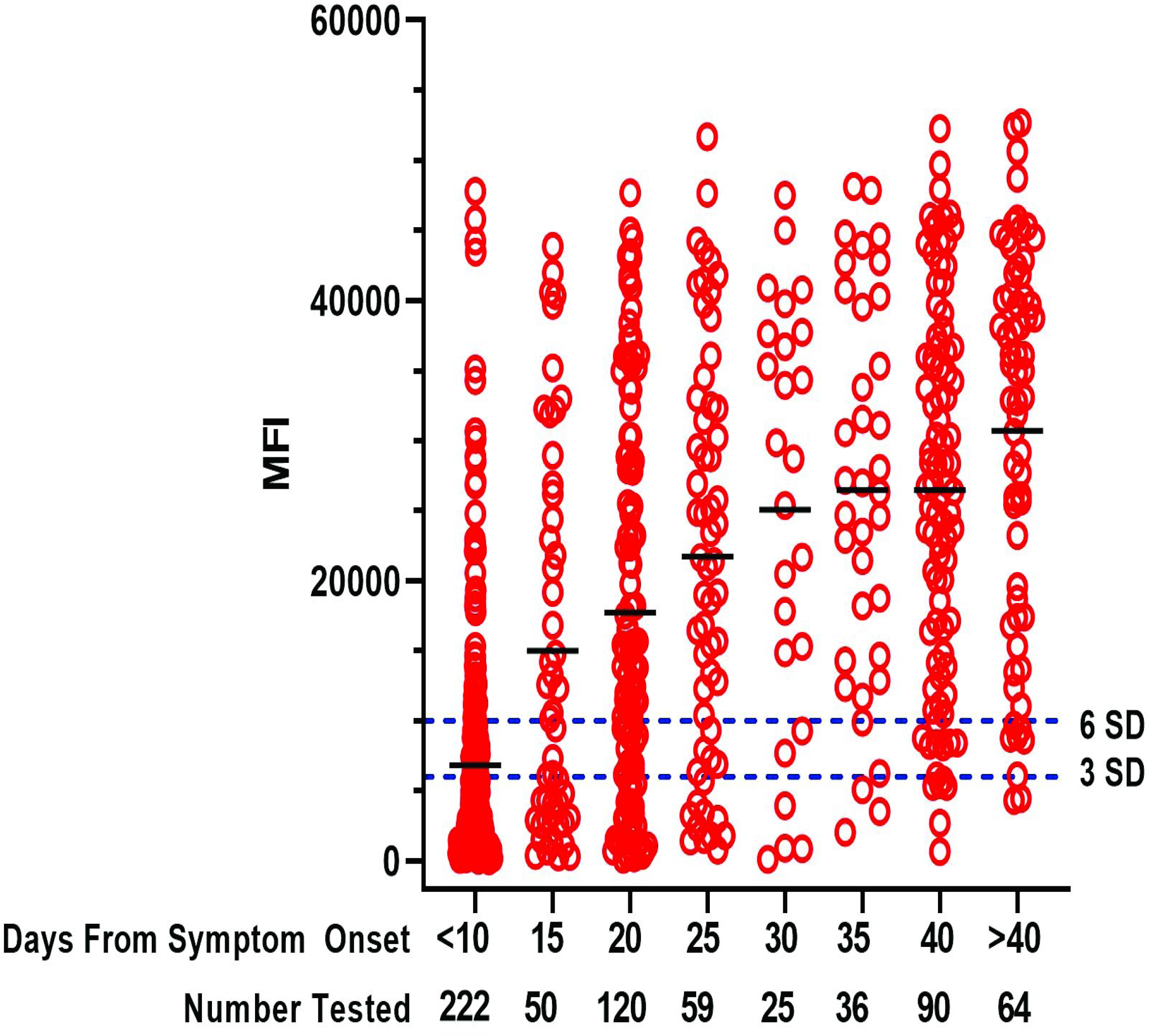
SARS-CoV-2 Ab Levels in Convalescent Sera. Serum specimens from COVID-19 convalescent patients (Mount Sinai and Westchester County) were tested at a 1:100 dilution in the MIA. MFI values, based on N antigen reactivity, from individual specimens are presented here grouped into 5-day blocks after symptom onset (except for <10 and >40). The cutoff at 6 SD (upper dotted line) and 3 SD lower dotted line) are shown.

In the MSH cohort, the titers of SARS-CoV-2 Abs were determined for those specimens that were positive for RBD binding at a greater than 1:50 dilution in the initial screen. Over the three-week course of the study, a higher proportion of sera showed increasingly higher titers, presumably reflecting maturation of the immune response. Supplementary Table 2 shows distribution of serum titers of samples received during the final 9 days of the study, when most individuals would be expected to be > one month post symptom onset (PSO). As indicated, 89.9% of the “screen positive” sera had SARS-CoV-2 titers that exceeded a 320 threshold. We noted that 10% of the Ab positive sera failed to meet a 320 titer threshold for strong Ab responses even three weeks from initial symptom onset. In sera that were collected from the WCDH cohort of convalescent individuals who were at least 21 days PSO, the MIA showed reactivity for that 83.2% (6 SD) or 90.6% (3 SD) of the specimens. A composite analysis of Ab levels, as indicated by differences in MIA signal intensities is shown in Figure 1.

**Table 2.** Relationship between SARS-CoV Neutralizing Abs and Onset of Symptoms.

| SARS-CoV-2 PRNT<sub>50</sub> Titers |  |  |  |  |  |  |  |
| --- | --- | --- | --- | --- | --- | --- | --- |
| Onset (Days) | PRNT tested | <80 | 80 | 160 | >320 | % Neutralizers at FDA minimal level | % Neutralizers at FDA recommended level |
| 11-15 | 12 | 5 | 4 | 2 | 1 | 58.3 | 25.0 |
| 16-20 | 58 | 28 | 12 | 7 | 11 | 51.7 | 31.0 |
| 21-25 | 54 | 6 | 8 | 17 | 23 | 88.9 | 74.1 |
| 26-30 | 53 | 2 | 10 | 7 | 34 | 96.2 | 77.4 |
| 31-35 | 41 | 0 | 3 | 8 | 30 | 100 | 92.7 |
| 36-40 | 34 | 1 | 5 | 7 | 21 | 97.1 | 82.4 |
| >40 | 48 | 3 | 12 | 10 | 23 | 93.8 | 68.8 |

| SARS-CoV-2 PRNT<sub>90</sub> Titers |  |  |  |  |  |  |  |
| --- | --- | --- | --- | --- | --- | --- | --- |
| Onset (Days) | PRNT tested | <80 | 80 | 160 | >320 | % Neutralizers at FDA minimal level | % Neutralizers at FDA recommended level |
| 11-15 | 12 | 11 | 0 | 0 | 1 | 8.3 | 8.3 |
| 16-20 | 59 | 42 | 8 | 2 | 7 | 28.8 | 15.3 |
| 21-25 | 54 | 19 | 15 | 13 | 7 | 64.8 | 37.0 |
| 26-30 | 53 | 12 | 16 | 13 | 12 | 77.4 | 47.2 |
| 31-35 | 41 | 7 | 12 | 10 | 12 | 82.9 | 53.7 |
| 36-40 | 34 | 13 | 13 | 3 | 5 | 61.8 | 23.5 |
| >40 | 48 | 28 | 10 | 5 | 5 | 41.7 | 20.8 |

### Relationship between antibody production and virus neutralizing antibody

A main protective function of anti-viral antibodies is to prevent infection by interfering with essential virus-host cell interactions such as receptor binding, virus entry, and membrane fusion [7] [21]. Neutralization can be measured by PRNT, which we used to determine the levels of neutralizing Abs in two cohorts of recovered COVID patients. All sera used were reactive in the NYS SARS-CoV MIA, and most specimens were tested using a multiplexed MIA that included the SARS-CoV-2 RBD along with the N protein. Our objective in this portion of the study was to identify those sera that met the recommended (160) or minimal (80) titers for convalescent plasma proposed by the FDA at both the 50% (PRNT50) and 90% (PRNT90) neutralization levels.

We examined the relationship between ELISA and PRNT titers using sera collected by MSH to determine whether a direct correlation could be made between Ab reactivity in the ELISA and neutralizing activity. Analysis of 159 sera showed that, as expected, there was a general trend toward more neutralization with higher Ab titers (Table 1). Approximately half (52.9%) of the samples had a ≥160 neutralizing titer PRNT50 level, including 84.1% of sera with an ELISA titer of 2880. However, only 50% of the sera at the highest ELISA Ab titer (2880) had neutralizing titers of 160 or greater when a more stringent (PRNT90) determination for neutralization was used. Figure 2 shows a graphical representation of these data, except that results are grouped to show the frequencies of specimens that met the minimal FDA recommended 80 and 160 titers for convalescent plasma use (“Medium”) in addition to frequencies of specimens that either failed to meet that recommendation (“Low”) or exceeded the recommended titers (“High”).

**Figure 2.**
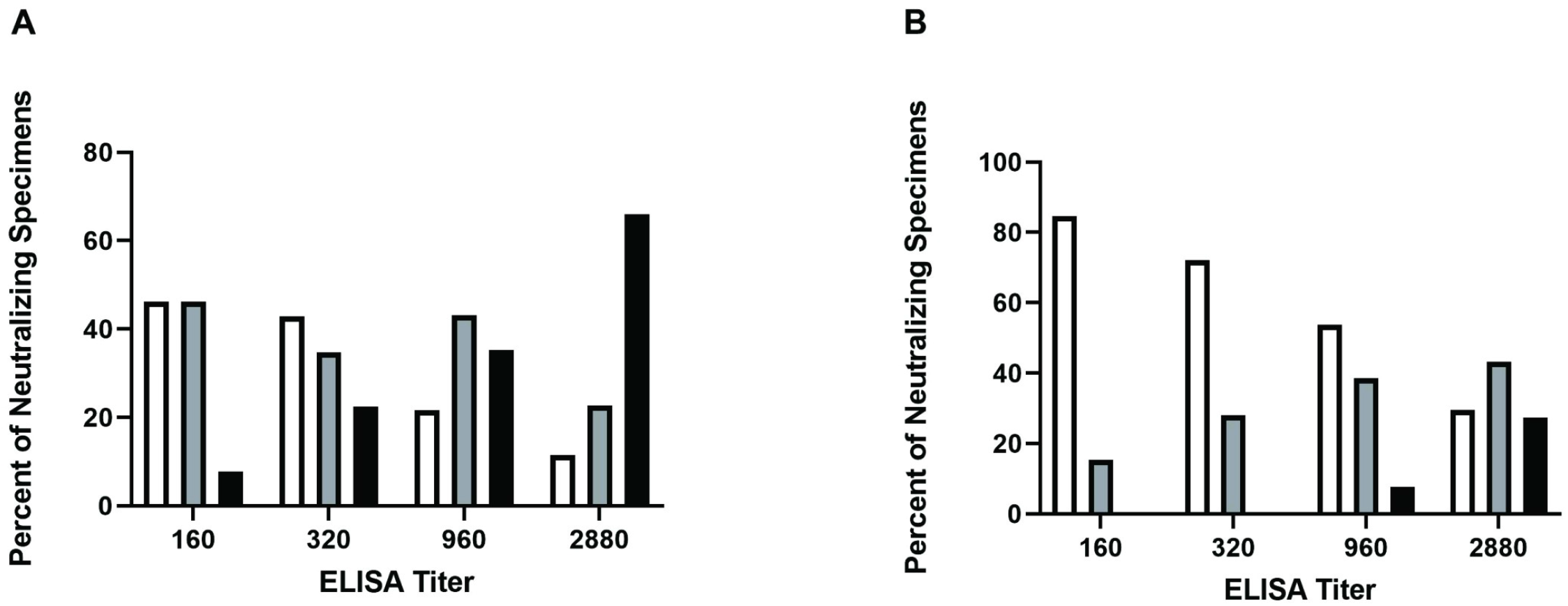
Relationship between SARS-CoV Neutralizing Abs and ELISA Titer. Serum specimens from COVID-19 convalescent patients that screened Ab positive to SARS-CoV-2 RBD, were titered by ELISA with respect to reactivity to SARS-CoV-2 whole spike protein. Sera with ELISA titers ≥160 were tested for their ability to neutralize SARS-CoV-2 infection of Vero E6 cells and the PRNT_50_ (A) and PRNT_90_ (B) titers were determined. Neutralizing activity is grouped according to titer: Low <80, *white bars*; Medium 80/160, *gray bars*; High ≥320, *black bars*.

In general, samples with high Ab titers by ELISA also had high MIA MFIs, consistent with overall agreement between the two Ab detection assays. Likewise, we found that specimens with higher amounts of Ab detected using the MIA, had higher levels of neutralizing Ab activity (Figure 3). Although more overall Ab led to more neutralizing Ab, we did not find a specific MFI value to be an absolute predictor of neutralizing activity. Specimens with similar Ab (MFI) levels could be found at each level of neutralization (Low, Medium, High), although their frequencies varied. In a smaller sampling, we substituted the RBD antigen for the N antigen in the MIA and, again, found no MFI value that predicted whether the specimen would be a strong neutralizer (data not shown).

**Figure 3.**
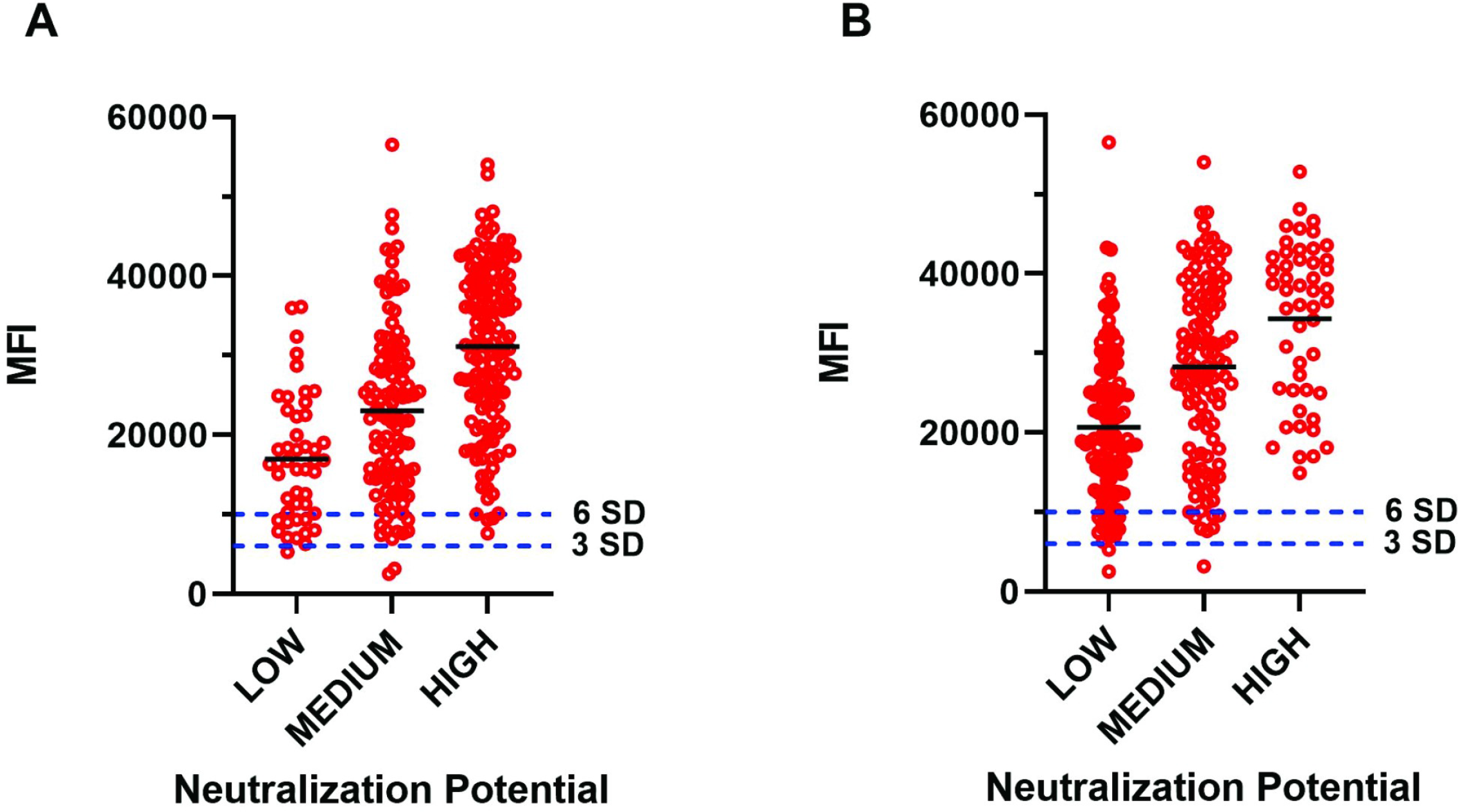
Relationship between SARS-CoV Neutralizing Abs and MIA MFI. Serum specimens from COVID-19 convalescent patients were assessed for N protein Ab levels by MIA and then their neutralizing capacity was measured at PRNT_50_ (A) and PRNT_90_ (B). Neutralizing activity is grouped according to titer: Low <80, Medium 80/160, High ≥320.

More individuals made higher levels of Ab as time from symptom onset increased, so we examined neutralization titers over different time periods PSO to explore whether more neutralizing Abs were produced after longer recovery periods. PRNT analysis of ∼300 sera from the combined cohorts demonstrated that, consistent with overall Ab production, neutralization activity also increased with time after symptom onset. The peak of neutralization in this study was at 31-35 days PSO (Table 2), where ∼93% of the sera had ≥ 160 PRNT50 titers, while ∼54% of the sera had ≥ 160 titers using the more stringent PRNT90 evaluation. Beyond 35 days, the number of sera which had high PRNT90 titers decreased significantly, with only ∼24% of the sera having ≥ 160 neutralizing titers (Table 2, Figure 4).

**Figure 4.**
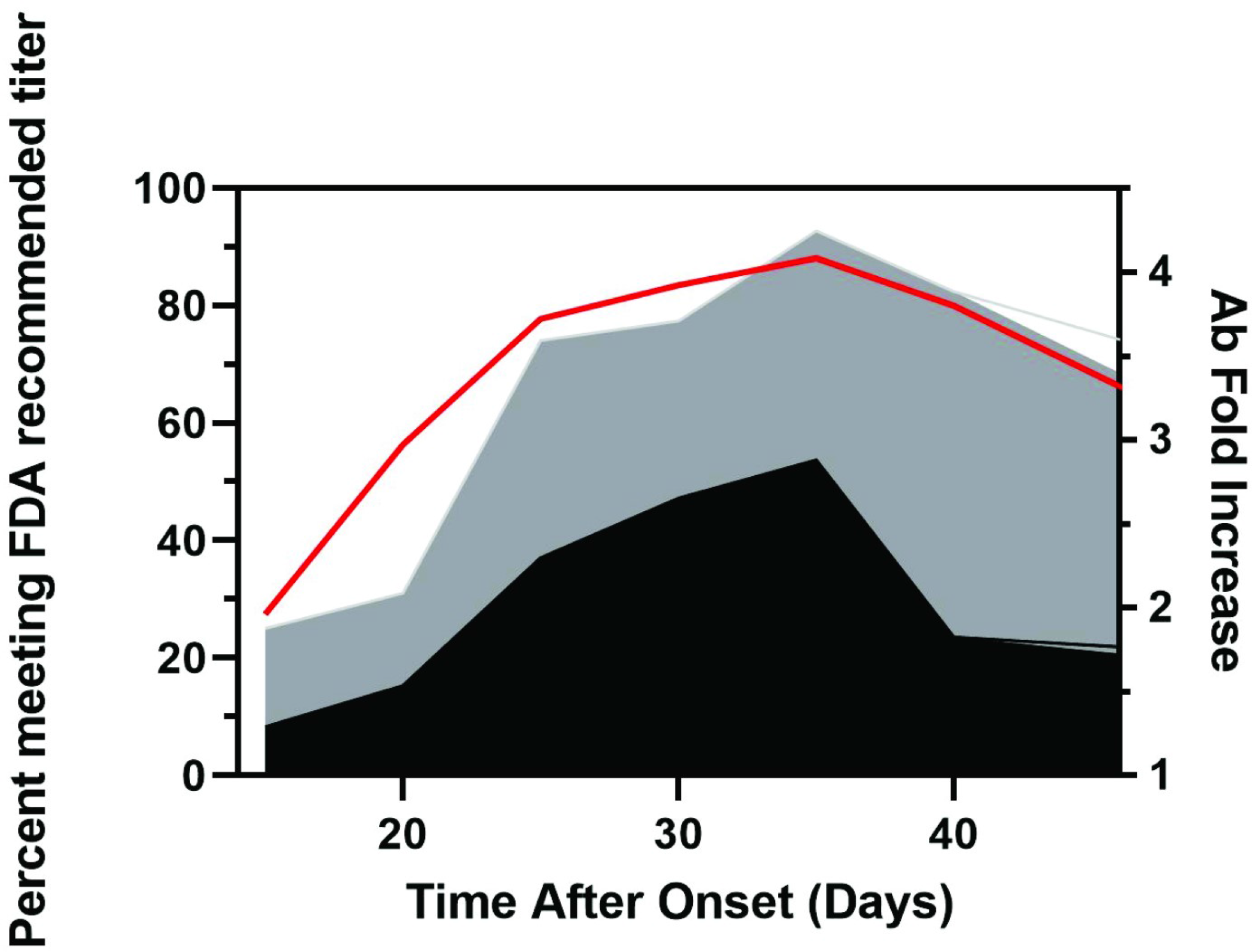
Relationship between SARS-CoV Neutralizing Abs and Onset of Symptoms. Graphical representation of the data shown in Figure 1 and Table 2. Shown on the left Y-Axis are the percentages of specimens that can neutralize 90% (**black**) or 50% (**gray**) of viral plaque formation with a titer of 160 or greater. Note that, since the last specimens collected were grouped as >40 days, 45 days was used as an arbitrary end point for graphing purposes. On the right axis is shown the fold increase of the mean levels of Ab production (**red line**), based on MFI’s from Figure 1.

## Discussion

At present, plasma from recovered COVID-19 patients for passive Ab transfer is being investigated as an experimental treatment for patients severely ill with COVID 19. Passive transfer is an old method that has seen recent use during infectious disease outbreaks to combat infections by pathogens, such H1N1 influenza virus, Ebola virus, and notably, MERS CoV and SARS CoV [22]. As a potential emergency therapy for COVID-19, at least two groups have reported compassionate use and some success in passive transfer of immune plasma into critically ill COVID-19 patients [23] [24]. Controlled trials are urgently needed to establish the parameters for effective therapy, but the success of this approach relies upon the presence of protective antibodies in the sera of recovered, presumably immune, individuals. The FDA has provided limited nonbinding recommendations for investigative COVID-19 convalescent plasma use based on current knowledge [11].

For viruses in general, a main advantage of Ab protection lies in its ability to block the interaction between the virus and its cellular receptor in the host, thereby preventing entry into the cell and viral replication [25]. Although the relationship between neutralizing Ab and immune protection from many virus infections is established, many questions remain to be answered about the levels of neutralizing Abs needed to confer immunity to SARS-CoV-2.

Surprisingly, we found that while most individuals made moderate to high levels of SARS-CoV-2 specific Ab, a smaller than expected number met the FDA recommended titers of neutralizing Abs [11], as determined by a PRNT. These results suggest that measurement of Ab neutralizing activity has significant potential to enhance the effectiveness of plasma therapy for COVID-19 through improved donor selection criteria. In this study most recovered COVID-19 individuals made detectable neutralizing Abs when measured using a PRNT50, but only half the sera with a 960 ELISA titer against the spike protein had a neutralizing titer of at least 160, the FDA recommended level of titer. While a 2880 ELISA titer was predictive of neutralizing activity at 160 with PRNT_50_, only 50% of these individuals reached the 160-titer recommendation at the PRNT90 level. The FDA has not recommended a specific assay or stringency level, and further studies are critically needed to define the optimal levels of neutralizing antibodies for plasma therapy [11].

Through the use of two cohorts, we also found that ELISA IgG results correlated well with the means of the MIA MFI values. However, we observed greater variation in SARS-CoV-2 reactivity among individuals in this study than we have in our previous experience using the MIA for Ab detection for other viruses [26, 27]. Whether the variation in antibody production that we observed here is unique to responses to SARS-CoV-2 needs further study.

Our results parallel the recent findings of Wu et al that approximately 30% of recovered COVID-19 patients, examined two weeks after discharge from hospitals, generated low titers of SARS-CoV2 specific neutralizing antibodies [28]. However, others have reported higher levels of correlation between antibody levels and neutralizing activity, including a smaller study that evaluated serial samples from 12 COVID-19 patients [29] and a larger study using a pseudo-typed virus neutralization assay [28]. A recent study using the MSH ELISA and an optimized and sensitive MN assay recently also found high IC_50_ neutralization titers and excellent correlation between ELISA and MN assay [30]. Our results indicate that high level Ab quantitation is a useful, but imperfect, guide to donor suitability, depending on the required level of neutralization. Additional factors, including the specific target antigen used to measure antibody levels, should also be considered.

As expected, we found that timing post infection had a major impact on the degree of neutralizing activity present. Our finding that SARS-CoV-2 neutralizing antibody responses are most abundant at 31-35 days PSO is similar to the >21-days PSO peak in neutralizing antibody titer observed for SARS-CoV-1 [31] but later than the two to three weeks PSO reported by others [28] [32] [29] [24]. The drop in neutralizing activity we observed beyond 35 days PSO raises the possibility that there is a relatively narrow window for choosing the “best” sera for treatment, particularly at the more stringent PRNT90 level of neutralization. It is critical to determine the level of neutralization required for effective plasma therapy as we further study this experimental treatment. In particular, the level of stringency required for neutralization must be defined, as this affects the interpretation of FDA recommendations regarding plasma titers. While the 1:160 neutralization recommendation from FDA may be higher than necessary, we note that the European Commission suggests using a minimum titer of 320 [33].

At present, the extent to which neutralizing Abs contribute to protection from reinfection by SARS-CoV-2 is not known and the use of either low stringency (PRNT50) or high stringency (PRNT90) evaluative methods might not reflect the true requirements for protection. For example, significant protection against influenza virus infection is observed at 40 titers [34] and protection from measles virus infection correlates with 120 PRNT_50_ titers [35]. However, the optimal level of neutralizing Abs for plasma therapy is likely to differ from that required for an individual’s own immune status, as donor plasma will be diluted as much as ten-fold upon transfer into a recipient and protection from viruses after infection or vaccination is not just limited to neutralizing antibodies [36]. The mechanisms for potential immunity to SARS-CoV-2 have yet to be determined. There are limited reports about the protective cellular immune response to SARS-CoV-2 [37], although there are numerous reports about cellular immunopathology and negative consequences for the disease [38]. Strong cellular immunity could more than compensate for low virus neutralizing capacity. A recent study testing an inactivated whole virion SARS-CoV-2 vaccine (PiCoVacc) in non-human primates reported low neutralizing titers post-vaccination but found evidence of protection from disease in animals challenged with SARS-CoV-2 three weeks after vaccination [39]. Thus, high neutralizing antibody titers may not be required for protection from reinfection with SARS-CoV-2.

While establishing the relationship between PRNT titers and efficacy of plasma therapy for treatment of COVID-19 will require the results of controlled trials, we think the data presented here point to the need to thoroughly characterize individual plasma used for passive Ab therapy. At a minimum, the time period between symptom onset and serum collection may need to be refined to prioritize establishing the precise PRNT titer of the sera prior to clinical use. Rapid neutralization assays that can be done in less restrictive biocontainment conditions and/or better-defined correlates for neutralizing activity are clearly needed for understanding and control of SARS-CoV-2 infection and should be prioritized for further study.

## Data Availability

All data referred to in the manuscript is freely available.

## Acknowledgments

The authors would like to acknowledge and thank members of the following Wadsworth Center laboratories for their expert technical assistance: Diagnostic Immunology (S. Brunt, S. Bush, K. Carson, J. Chan, A. Cukrovany, V. Demarest, A. Furuya, K. Howard, D. Hunt, N. Jones, J. Kenneally, C. Koetzner, M. Marchewka, R. Stone, C. Walsh, J. Yates). We would like to acknowledge the Wadsworth Center core facilities that contributed to this work: WC Tissue Culture and Media. We would like to acknowledge the following members of the Westchester County Department of Health for their many contributions to the success of this project: L. Smittle, R. Recchia, J. Li, T. Peer, J. Falasca, E. Cestone, and L. Goldsmith. We would like to thank Dr. R. Amler for helpful discussions and for critical reading of this manuscript

## Competing interests

MSH is in the process of licensing assays to commercial entities based on the assays described here and has filed for patent protection.

## Abbreviations used

FDA: Food and Drug Administration
PRNT: plaque reduction neutralization test
PSO: post-symptom onset
COVID-19: Coronavirus Disease 2019
SARS-CoV-2: Severe Acute Respiratory Syndrome coronavirus 2
Ab: antibody
MN: microneutralization
MSH: Mount Sinai Hospital
WCDH: Westchester County Department of Health
ELISA: enzyme-linked immunosorbent assay
MIA: microsphere immunoassay
N: nucleocapsid protein
RBD: receptor-binding domain of the spike protein

## Notes

Competing interests **–** Mount Sinai Hospital is in the process of licensing assays to commercial entities based on the assays described here and has filed for patent protection.

This work was supported by the New York State Department of Health

### Competing Interest Statement

Mount Sinai Hospital is in the process of licensing assays to commercial entities based on the assays described here and has filed for patent protection.

### Funding Statement

Testing at the Wadsworth Center was supported by internal funding at the New York State Department of Health. Mount Sinai Hospital is a CLIA laboratory and all the testing for patient specimens was supported by reimbursement from insurance policies.

